# Community Threat, Positive Parenting, and Accelerated Epigenetic Aging: Longitudinal Links from Childhood to Adolescence

**DOI:** 10.1101/2024.12.23.24319484

**Authors:** Georgette Metrailer, Karina Tavares, Mikayla Ver Pault, Adamari Lopez, Shane Denherder, Evelyn Hernandez Valencia, Karissa DiMarzio, April Highlander, Sarah M. Merrill, Darlynn M. Rojo-Wissar, Justin Parent

## Abstract

Early life adversity (ELA) has been linked to accelerated epigenetic aging. While positive parenting is hypothesized to buffer the effects of ELA, its role in mitigating epigenetic age acceleration remains unclear. Data from 2,039 children (49.7% female; 46.7% Black, 26.5% Hispanic, 19% White non-Hispanic) in the Future of Families and Child Wellbeing Study were included. Home threat, community threat, and parenting were measured from ages 3 to 9 (2001– 2010). Epigenetic age acceleration was assessed at ages 9 and 15 (2007–2017). Positive parenting reduced the pace of epigenetic aging in contexts of low, but not high, community threat (β = .026, p = .039). Interventions across multiple socioecological systems may be necessary to prevent the biological embedding of ELA.

---

Early life adversity (ELA) encompasses a wide array of challenging environmental experiences during childhood, including exposure to violence, abuse, neglect, and chronic poverty (Madigan et al., 2023). These experiences often diverge significantly from typical developmental environments, demanding substantial adaptive responses from affected children (Humphreys & Zeanah, 2015; Merrill, Konwar, et al., 2024; Nelson III & Gabard-Durnam, 2020). In a meta-analysis of 206 studies, Madigan and colleagues (2023) showed that 60.1% of adults in 22 countries report at least one adverse childhood experience, with 16.1% reporting four or more. Additionally, research has consistently demonstrated that having a history of ELA is closely linked to multiple mental (LeMoult et al., 2020) and physical (Grummitt et al., 2021) health problems, highlighting the urgent need for further investigation into factors that mitigate the negative outcomes associated with ELA.

## Early Life Adversity and Epigenetic Aging

ELA is associated with long-term health outcomes in part due to the physiological impact of stress, which can become biologically embedded (Aristizabal et al., 2020). Biological embedding refers to environmentally induced alterations in physiological systems that may result in enduring biological changes (Aristizabal et al., 2020). DNA methylation, a key mechanism of biological embedding (Aristizabal et al., 2020), involves the addition of a methyl group to DNA, typically at a cytosine base, followed by a guanine base (CpGs). This epigenetic modification can influence gene expression without altering the underlying DNA sequence (Aristizabal et al., 2020). Crucial for processes like cell differentiation and development, DNA methylation patterns also shift predictably with cellular aging, thereby altering cellular behavior and responses to environmental stimuli throughout life (Horvath, 2013). Interestingly, a recent study indicated that ELA is associated with widespread alterations in DNA methylation as early as infancy and preschool age, highlighting early childhood as a particularly sensitive window for epigenetic regulation (Lussier et al., 2023). In addition, adolescence has emerged as a second sensitive period for epigenetic remodeling and the consolidation of stress-related physiological changes– particularly within immune function and emotional regulation systems (Suglia et al., 2018). Therefore, assessing DNA methylation during adolescence may allow for the early detection of lasting biological effects of ELA, and may serve as a critical period for intervention before the emergence of negative health outcomes in adulthood.

Researchers can estimate an individual’s biological or cellular age by analyzing specific DNA methylation sites across the genome, providing valuable insights into how early environmental exposures influence biological processes (Horvath, 2013). Epigenetic age acceleration occurs when biological age, as estimated with DNA methylation, differs from chronological age with increased acceleration indicating advanced biological aging relative to one’s actual age (Horvath, 2013). Two widely used epigenetic clocks in pediatric populations are the Pediatric Buccal Epigenetic Clock (PedBE) and the DunedinPACE. These clocks were developed for different purposes and capture distinct aspects of biological change. The PedBE is a specialized tool designed to estimate DNA methylation age in children’s oral samples with the primary goal of measuring developmental timing rather than disease risk (Fang et al., 2023). The PedBE is highly correlated with chronological age and serves as an indicator of developmental success or deviations from typical development trajectories in children (Kusters & Horvath, 2024). For example, recent studies have used the PedBE to detect behavioral changes in response to early interventions such as Parent-Child Interaction Therapy, thereby highlighting its sensitivity to behaviorally relevant developmental shifts (Sullivan et al., 2023).

In contrast, the DunedinPACE measures the pace of biological aging by assessing longitudinal changes across seven organ systems—cardiovascular, metabolic, renal, hepatic, immune, dental, and pulmonary (Belsky et al., 2022). It was specifically developed to capture multisystem physiological decline and has shown strong associations with morbidity, mortality, and functional decline in adulthood (Belsky et al., 2022) across multiple diverse ethnic backgrounds such as Native Americans, African Americans, and Hispanics (Kusters & Horvath, 2024). While both clocks are significantly correlated with life– and health-span outcomes, they were developed for different purposes and represent different aspects of biological change across the lifespan. Therefore, using multiple clocks may allow for a more comprehensive examination of adversity affects overall health by capturing both deviations in normative developmental timing (PedBE) and the acceleration of multisystem biological aging (DunedinPACE). This multi-clock approach aligns with recent recommendations to use complementary aging biomarkers to better understand how social and environmental exposures shape diverse biological pathways, developmental outcomes, and health trajectories (Kusters & Horvath, 2024).

## Theoretical Models of Early Life Adversity

Life History Theory (LHT) offers one evolutionary-developmental framework for understanding how ELA may shape long-term biological and physical health outcomes. LHT suggests that exposure to harsh, unpredictable environments may promote adaptive calibrations to biological systems, favoring accelerated aging or development and shortened investment in long-term health to enhance survival and reproductive success under conditions of moderate to extreme adversity (Ellis et al., 2009). From this perspective, earlier biological aging could reflect an adaptation to environmental demands rather than solely a marker of dysfunction. However, more recent dimensional models of adversity have further clarified how distinct types of ELA, such as threat and deprivation, influence specific developmental and mechanistic processes (e.g., cognitive performance) (McLaughlin et al., 2014). In addition, considering the environmental contexts in which adversity occurs—such as parenting quality and community well-being—may provide a more complete understanding of how adversity becomes biologically embedded during development.

While LHT highlights the importance of environmental harshness and unpredictability, more recent dimensional models have sought to specify distinct types of adversity and their differential effects on developmental pathways. Emerging frameworks, such as the Dimensional Model of Adversity and Psychopathology, address this need by categorizing ELA into two core dimensions: threat (e.g., exposure to violence or abuse) and deprivation (e.g., neglect, lack of cognitive or social stimulation) (McLaughlin et al., 2014). This model posits that different types of adversity influence distinct neurobiological and psychosocial mechanisms, ultimately leading to divergent developmental outcomes (McLaughlin et al., 2014). A growing body of research supports this distinction, demonstrating unique associations between these dimensions and the physiological processes implicated in biological embedding (Colich et al., 2020; McLaughlin et al., 2014). For example, recent studies have shown that threat-related adversity—particularly exposure to violence or abuse—is more consistently linked with accelerated epigenetic aging than deprivation-related experiences (Hogan et al., 2024; Chang et al., 2024).

Investigating the specific dimensions of ELA has been essential for uncovering distinct developmental outcomes, but considering the contexts in which adversity occurs may further clarify how these outcomes vary depending on the environmental systems through which adversity is experienced. Bronfenbrenner’s Bioecological Model (Bronfenbrenner & Ceci, 1994) offers a useful framework for understanding how environmental influences shape child development through dynamic, reciprocal interactions across multiple nested systems. This model suggests that development is driven by proximal processes that occur within the immediate microsystem, such as parent-child interactions, while broader contextual forces–like neighborhood characteristics–operate through the ecosystem therefore influencing the child both indirectly and directly. The mesosystem reflects the interplay between these levels, capturing how experiences in one setting (e.g., caregiving) can interact with conditions in another (e.g., the neighborhood) to influence developmental outcomes. This framework highlights the importance of examining not only the direct caregiving relationships but also the surrounding environments in which children grow, as both “layers” or environmental contexts contribute to biological and psychological development. Hopefully, by continuing to examine outcomes related to specific dimensions of ELA and the specific environments in which they may occur, researchers can better identify targeted pathways linking early experiences to later psychopathology, health disparities, and potentially psychological resilience. Further, by delineating between different environmental contexts, research may be able to pinpoint specific environmental contexts in which interventions may be the most beneficial.

## The Importance of Parenting

Parenting behaviors and the quality of parent-child relationships have emerged as significant factors that may moderate the effect of ELA on epigenetic aging trajectories (Brody et al., 2016; Sullivan et al., 2023). Positive parenting practices—characterized by warmth, sensitivity, and responsiveness—are critical in shaping children’s developmental experiences and influencing epigenetic regulation (Brody et al., 2016; Sullivan et al., 2023). In particular, supportive caregiving during early childhood is essential for calibrating the developing stress response system, with long-term implications for biological and psychological functioning. This period of heightened neurobiological plasticity represents a window in which caregiving may serve as a powerful buffer, shaping the trajectory of biological embedding across the lifespan (Brody et al., 2016; Sullivan et al., 2023). For example, threat-related adversity has been linked to alterations in biological and neurodevelopmental processes, including accelerated epigenetic aging (Colich et al., 2020); however, other studies have shown that nurturing parenting practices have the potential to mitigate these detrimental effects (Brody et al., 2016; Sullivan et al., 2023). Merrill et al. (2024) demonstrated that children who participated in internet-based parent-child interaction therapy (PCIT)—which promotes positive parenting strategies to manage child behavior—exhibited a slower pace of epigenetic aging compared to those in the control condition. Similarly, Sullivan et al. (2024) found that children involved in child-parent psychotherapy, an evidence-based dyadic psychosocial intervention, showed reduced epigenetic age acceleration. These findings, alongside other evidence that positive parenting can buffer the effects of adversity on epigenetic aging (Brody et al., 2016; Sullivan et al., 2023), underscore the potential for early caregiving to serve as a modifiable protective factor in the context of ELA.

## Current Study

To date, most studies examining the link between ELA and accelerated epigenetic aging have focused on a single developmental stage or concentrated on epigenetic outcomes specifically in adulthood. Additionally, the extant literature has predominantly evaluated the negative impacts of stress or negative parenting behavior on biological embedding, with limited exploration and attention given to protective factors or resilience trajectories. There is a critical need for social epigenetic research that spans multiple developmental stages, integrates multi-level analyses, and investigates potential protective factors. These approaches may inform more robust resilience models and prevention programs that move beyond deficit-based frameworks to better support youth who have experienced ELA. To address these gaps, the current study leverages data from the Future of Families and Child Wellbeing Study to prospectively examine the moderating role of observed positive parenting practices in longitudinal associations between childhood ELA and adolescent epigenetic aging (see Figure 1 for a conceptual model).

**Figure 1.**
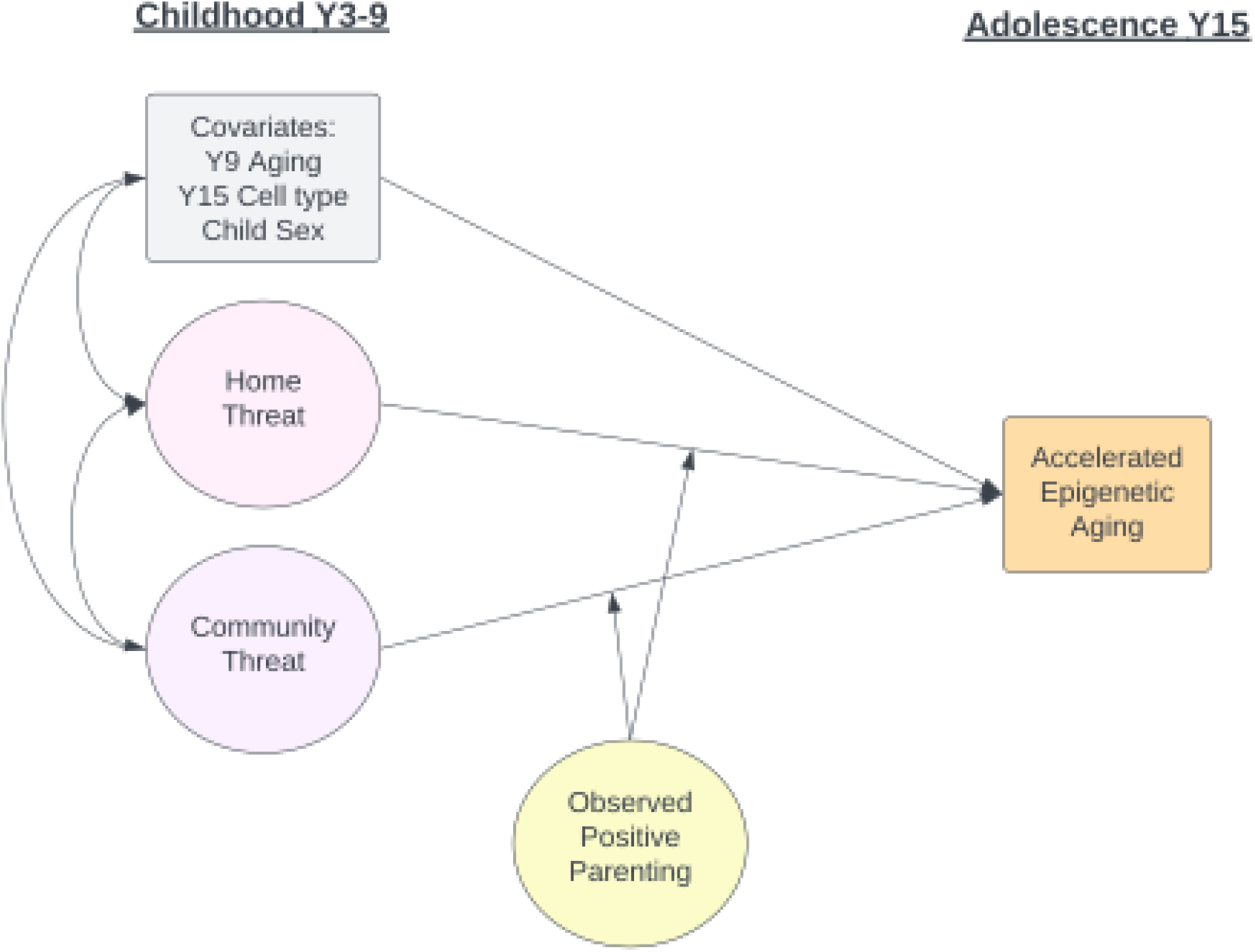
Conceptual model of the moderating effect of parenting on the link between childhood threat adversity and adolescent epigenetic age acceleration.

Building on prior findings linking threat-based ELA to accelerated epigenetic aging (Chang et al., 2024; Del Toro et al., 2024; Hogan et al., 2024), this study focused on threat-based ELA experienced between birth and age 9 occurring in both home (i.e., physical and/or emotional abuse) and community (i.e., crime and/or exposure to violence) settings. Further, to strengthen developmental inferences, this study assessed ELA and parenting prior to the measurement of epigenetic aging in adolescence. This temporal ordering enhances the study’s ability to examine longitudinal associations and potential mechanisms of biological embedding. Lastly, based on the previous literature outlined, it was hypothesized that observed positive parenting practices at ages 3, 5, and 9 will buffer the detrimental effects of threat-based ELA on accelerated epigenetic aging in adolescence.

## Method

### Participants

Data were drawn from the Future of Families and Child Wellbeing Study, an ongoing longitudinal study of 4,898 families from 20 large cities (population 200,000) across the United States (Reichman et al., 2001). Families were recruited at the child’s birth (1998-2000), and non-marital births were oversampled at a rate of 3:1, resulting in a sample enriched with economically disadvantaged families. The present study examined a subset of children (*n* = 2,039) from the larger sample, who were selected for having complete epigenetic data at age 9 or 15 years (2007-2017). Data for the present study were collected at birth and when the children were 3, 5, 9, and 15 years of age. See Table 1 for sample demographics. Sex was reported by the mother at birth, and the child’s race was self-reported at age 15 years.

**Table 1.**
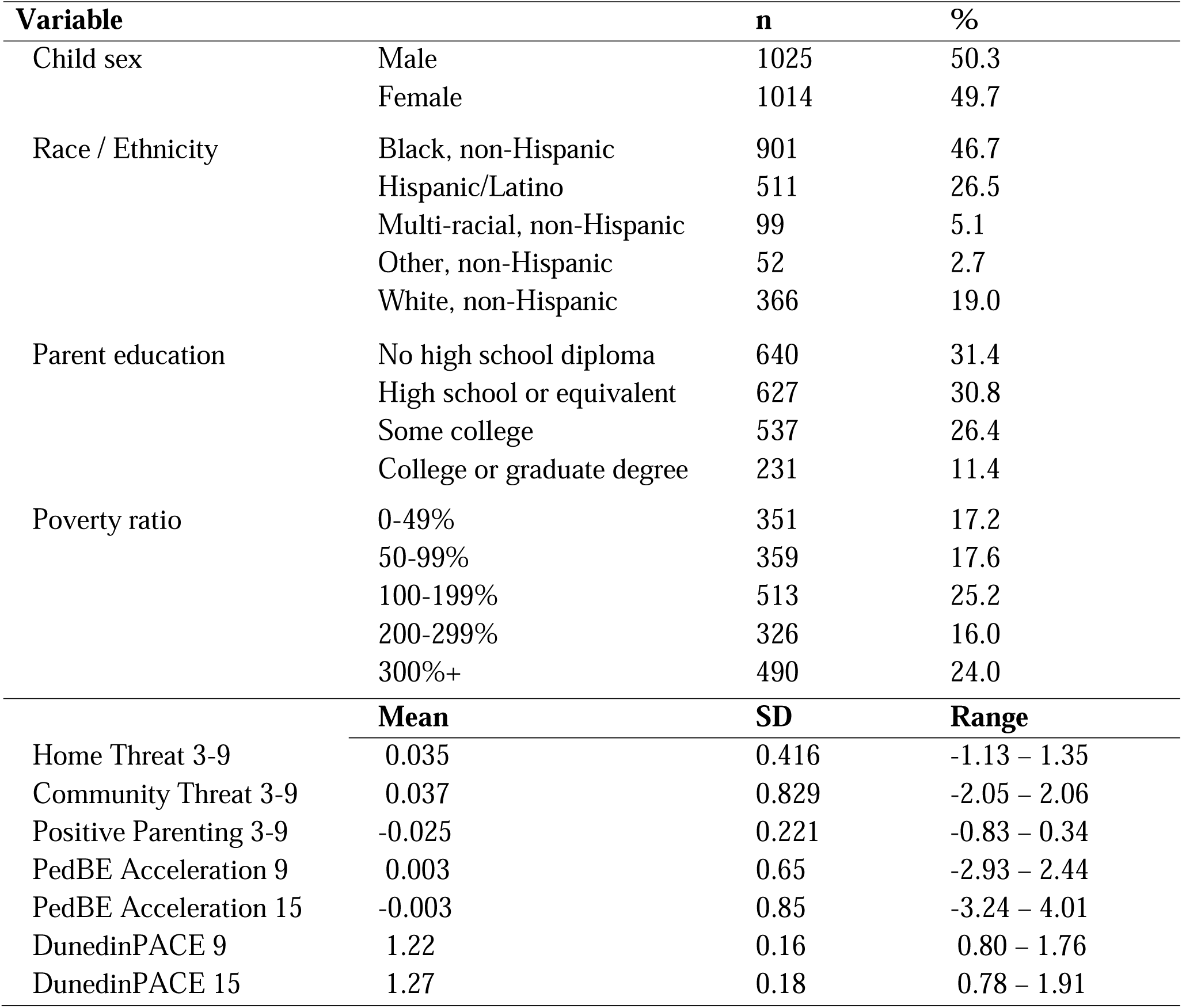
Sociodemographic and study variable descriptives.

### Procedure

Data for the present study were collected when the focal children were 3 (2001-2003), 5 (2003-2006), 9 (2007-2010), and 15 (2014-2017) years of age via in-home assessments. Children’s saliva samples were also collected during data collection at ages 9 and 15. In all, 86% of children provided saliva samples at age 9, as did 71% of teens at age 15. Data collection and study procedures were overseen by the Princeton University Institutional Review Board.

### Measures

#### Home Threat

[Author information redacted for review] and colleagues (YEAR) constructed a latent variable representing home threat using data collected at ages 3, 5, and 9 through the Parent–Child Conflict Tactics Scale (CTS-PC) (Straus et al., 1998). Home threat was defined as exposure to physical and emotional abuse by a primary caregiver. Primary caregivers reported the frequency of physical abuse behaviors (e.g., “spanked on the bottom with a bare hand”) and emotional abuse (e.g., “shouted, yelled, or screamed”) within the past year. Each category included three items, rated on a 7-point Likert scale ranging from “0 – never happened” to “6 – more than 20 times.” Higher scores indicated greater exposure to threatening experiences in the home. A hierarchical latent variable model was estimated using the Weighted Least Squares Mean and Variance adjusted estimator (WLSMV) with the first order including separate latent home threat variables for each age, and then a second-order factor representing a generalized construct of home threat that spans early childhood. Factor loadings were all above.60 for first-order factors and ranged from .68 to .87 for the second-order factor. Although the chi-square test was significant, χ²(51) = 224.396, p < .001, which is common in large samples, the other fit indices indicated good model fit: RMSEA = 0.030, 90% CI [0.026, 0.034]; CFI = 0.980; TLI = 0.974; and SRMR = 0.054. We then extracted the second-order factor score to be used as an observed variable in primary analyses. Complete details of the model are provided in the Supplementary Appendix (Tables S1 & S2 and Figure S1).

#### Community Threat

A latent variable was created to represent community-based threat across ages 3, 5, and 9, obtained via the National Archive of Criminal Justice Data’s Uniform Crime Reports. The data consisted of county-level crime rate data for the location of the focal child’s primary caregiver during data collection at ages 3, 5, and 9. The measure of community threat represents exposure to violence in the child’s neighborhood, calculated as the sum of violent crime instances per capita (murder, rape, robbery, and aggravated assault) and property crime instances per capita (burglary, larceny, motor vehicle theft, and arson). A single latent variable for community threat across early childhood was created with an indicator for each age that had factor loadings all above .76 and an excellent model fit. We then extracted the second-order factor score to be used as an observed variable in primary analyses. Complete details of the model are provided in the Supplementary Appendix (Tables S1 & S2 and Figure S1).

#### Parenting Practices

Positive Parenting at ages 3, 5, and 9 was measured via the Home Observation for Measurement of the Environment (HOME) Scale by a trained member of the FFCWS team (Bradley & Caldwell, 1984). The HOME Scale captures data regarding the caring environment in which the focal child was raised and has demonstrated strong interrater reliability and internal consistency (Bradley & Caldwell, 1984). The observer coded each item on a binary scale of “0 – did not occur” to “1 – did occur.” The HOME Scale was previously shown to be associated with prosocial (Blume et al., 2022) and externalizing behavior (Flannery et al., 2023) in the FFCWS. Similar to the above threat factors, a hierarchical latent variable model was estimated with first-order factors at each age and then a second-order factor capturing positive parenting across all three waves. Model fit was excellent, χ² (51) = 224.40, RMSEA = .030 [0.026, .034], CFI = .98, SRMR = 0.054, with standardized factor loadings within wave ranging from .68 to .94 and the second order factor loading being .49 for age 3, .70 for age 5, and .37 for age 9. We then extracted the second-order factor score to be used as an observed variable in primary analyses. Higher scores on the latent positive parenting variable represent higher levels of observed parent praise, positivity, warmth, and encouragement/support across the 3, 5, and 9-year waves. Complete details of the model are provided in the Supplementary Appendix (Tables S1 & S3 and Figure S2).

### Biological Markers

#### Epigenetic Aging

Saliva samples were collected by Westat Inc., the subcontractor for the FFCWS survey, during in-home visits when children were 9 and 15 years old. Trained interviewers used the Oragene® DNA Self-Collection kits (OGR-500; DNA Genotek Inc.) for sample collection. A total of 3,945 samples underwent analysis with methylation arrays (Infinium Human Methylation 450K and Infinium Methylation EPIC; Illumina), following the manufacturer’s guidelines. Quality control, conducted using the ENmix R package procedure, excluded samples flagged for outlier methylation or bisulfite conversion values, as well as those with sex mismatches between recorded data and methylation predictions. Cell-type proportions in saliva were estimated using the Houseman algorithm, implemented through the *estimateLC* function in the *ewastools* package, and referenced against the children’s saliva panel (Middleton et al., 2022). Epigenetic aging was estimated using two DNA methylation-based approaches, outlined below.

#### DunedinPACE

The DunedinPACE pace of aging (Belsky et al., 2022), previously applied to pediatric saliva samples (Merrill et al., 2024; Hogan et al., 2024; Raffington et al., 2021; Raffington et al., 2023), was utilized to assess epigenetic aging. This measure produces a value of one when epigenetic and chronological age align, while values greater than one indicate accelerated epigenetic aging relative to chronological age. Unlike traditional age-based epigenetic estimators, this biomarker was designed to reflect the physiological processes underpinning healthy biological aging (Belsky et al., 2022). Its development involved training on longitudinal data from the Dunedin Study, which tracked within-individual changes in 19 markers of organ-system integrity over two decades. Among the two DunedinPACE measures available from the FFCWS, this study employed the latest version (poam45).

#### PedBE Epigenetic Age Acceleration

Estimates of DNA methylation age acceleration in children were calculated using the Pediatric Buccal Epigenetic Clock (PedBE) (McEwen et al., 2020). This clock, specifically trained in oral tissue, estimates biological age in children with a margin of error under 4 months by analyzing 95 epigenome sites. PedBE age acceleration was determined as the residuals from a linear mixed-effects model, where predicted PedBE age was regressed on chronological age using maximum likelihood estimation. The model accounted for predicted buccal epithelial cell proportion (following the tool’s recommended procedure; McEwen et al., 2020) and included a random effect for individual participants (both ages 9 and 15 were included). These analyses were conducted in R (version 4.3.1) using the nlme package. Buccal epithelial cell proportions, estimated with the EpiDISH package, were included in the calculation of epigenetic age acceleration due to their established association with age.

### Statistical Analysis Plan

A path analysis model was conducted in Mplus using Full Information Maximum Likelihood (FIML) to handle missing data, with maximum likelihood estimation and robust standard errors (MLR) applied to account for potential non-normality. Model fit was assessed using the following criteria: chi-square, χ2: p > 0.05 excellent, comparative fit index (CFI; > 0.90 acceptable, > 0.95 excellent), root mean square error of approximation (RMSEA; < 0.08 acceptable, < 0.05 excellent), and the standardized root mean square residual (SRMR; < 0.08 acceptable, < 0.05 excellent). Two models were run with a similar set of predictors, with the outcome differing between the two epigenetic aging outcomes. For each model, covariates were child sex, buccal epithelial cell (BEC) proportion, and age 9 levels of the epigenetic outcome. Following best practices for salivary epigenetic age estimation in pediatric samples (Merrill et al., 2025), we included BEC proportion at the stage of calculating accelerated aging and as a covariate. This dual adjustment reduces variance due to cellular heterogeneity while addressing potential confounding due to developmental or environmental correlates of cell-type composition. For each model, the core predictors were home threat, community threat, observed positive parenting, and the two interactions: home threat by positive parenting and community threat by positive parenting. Positive parenting, home threat, and community threat were included in the model as observed variables based on the extracted factor scores described above. The interaction term involved multiplying observed scores. Simple slopes for high (+1 SD), mean, and low (–1 SD) levels of positive parenting were estimated and plotted to interpret any significant interaction effect.

### Exploratory vs. Confirmatory Statement

The present analyses were not preregistered. This study represents a combination of confirmatory and exploratory elements. The primary hypotheses—that positive parenting would buffer the effects of threat-based adversity on accelerated epigenetic aging—were specified a priori based on prior literature and theoretical models. However, some aspects of the analytic plan (e.g., differential effects across distinct epigenetic aging measures, examination of specific interaction patterns) were exploratory in nature, as the field currently lacks clear consensus about which biological pathways or metrics best capture these effects in childhood and adolescence. Thus, the findings should be interpreted within this mixed confirmatory-exploratory framework.

## Results

### Preliminary results

Descriptives for epigenetic aging outcomes across waves suggest the sample, on average, showed no acceleration on PedBE. In contrast, they showed a faster-than-expected aging pace on the DunedinPACE, though there was substantial variability in each outcome (see Table 1). Bivariate correlations are presented in Figure 1. <<<Insert Table 1 here>>> <<<Insert Figure 1 here>>> Community, but not home, threat adversity was positively correlated with accelerated epigenetic aging, whereas positive parenting was negatively correlated with the pace of aging. BEC cell type was correlated with each epigenetic outcome, and child sex was associated with the pace of aging such that girls had a faster pace of aging. Overall, associations support the inclusion of the covariates and proceeding to the primary models.

### Primary results

Complete results are detailed in Table 2, while the simplified conceptual moderation model is illustrated in Figure 1. <<<Insert Table 2 here>>> The model fit for the pace of aging model was excellent, with χ² (6) = 5.999, p = 0.423, RMSEA = .000 [.000, .029], CFI = 1.0, SRMR = 0.009. Similarly, the model fit for accelerated PedBE epigenetic age was also excellent, χ² (6) = 5.815, p = 0.444, RMSEA = .000 [.000, .028], CFI = 1.0, SRMR = 0.008. For the Dunedin pace of aging outcome, higher levels of childhood community, but not home, threat predicted a faster pace of aging in adolescence over and above the effect of child sex, cell type, and age 9 pace of aging. Further, higher levels of observed positive parenting in childhood predicted a slower pace of epigenetic aging in adolescence. Importantly, the interaction between community threat and positive parenting was significant, while the interaction with home threat was not. For the PedBE accelerated epigenetic aging outcome, higher levels of childhood community and home threat predicted adolescent accelerated epigenetic aging over and above the effect of child sex, cell type, and age 9 PedBE. However, observed positive parenting was not related to PedBE acceleration, nor was either interaction significant. Across models, we examined sensitivity analyses by adding the family poverty ratio as an additional covariate to better understand the unique effects of threat-based adversity over socioeconomic factors. For both outcomes, the results remained consistent, with similar main effects and a significant interaction for the pace of aging but not for PedBE-accelerated epigenetic aging.

**Table 2.** Primary Path Analysis Model Results.

| | b | 95% CI | $\beta$ | *p* |
| --- | --- | --- | --- | --- |
| **DV: Dunedin PACE Y15** |  |  |  |  |
| Community Threat | 0.012 | 0.006 – 0.017 | 0.054 | .000 |
| Home Threat | 0.007 | -0.004 – 0.019 | 0.017 | .214 |
| Positive Parenting | -0.040 | -0.061 – -0.018 | -0.049 | .000 |
| BEC Y15 | 0.809 | 0.772 – 0.846 | 0.735 | .000 |
| Dunedin PACE Y 9 | 0.237 | 0.206 – 0.267 | 0.211 | .000 |
| Child Sex | 0.055 | 0.045 – 0.064 | 0.152 | .000 |
| CommunityT * Parenting | 0.025 | 0.001 – 0.048 | 0.026 | .039 |
| HomeT * Parenting | 0.025 | -0.023 – 0.074 | 0.014 | .308 |
| **DV: PedBE EAA Y15** |  |  |  |  |
| Community Threat | 0.074 | 0.040 – 0.109 | 0.073 | .000 |
| Home Threat | 0.114 | 0.043 – 0.185 | 0.056 | .002 |
| Positive Parenting | -0.036 | -0.174 – 0.102 | -0.009 | .606 |
| BEC Y15 | 1.420 | 1.184 – 1.618 | 0.272 | .000 |
| PedBE EAA Y 9 | -0.576 | -0.636 – -0.515 | -0.442 | .000 |
| Child Sex | 0.080 | 0.019 – 0.141 | 0.047 | .010 |
| CommunityT * Parenting | -0.089 | -0.256 – 0.077 | -0.020 | .293 |
| HomeT * Parenting | 0.077 | -0.248 – 0.402 | -0.020 | .642 |

Note. BEC = buccal epithelial cell proportion; Y9 = year 9; Y15 = year 15; CommunityT = community threat; HomeT = home threat. $\beta$ = standardized beta coefficient

Figure 3 illustrates the simple slopes of the interaction between community threat and positive parenting in childhood on the adolescent pace of epigenetic aging. <<<Insert Figure 3 here>>> Levels of plotted slopes were for +/− 1 SD. Results indicate that low levels of observed positive parenting (–1 SD) mitigate the influence of community-level adversity, leading to an accelerated pace of aging regardless of the level of community threat (b = .006, p = .170). In contrast, when positive parenting was average (b = .012, p < .001) or high (+1SD) (b = .018, p < .001), community threat had a longitudinal effect on the pace of aging, with greater threat associated with faster aging. Furthermore, findings suggest that high levels (+1SD) of community threat (b = –.015, p = .354) weaken the effect of positive parenting on pace of aging. In other words, an accelerated pace of aging occurs when either positive parenting is low or community threat is high, regardless of the other factor. Adolescents demonstrated the slowest epigenetic pace of aging when exposed to high levels of positive parenting and low levels of community threat during childhood.

**Figure 2.**
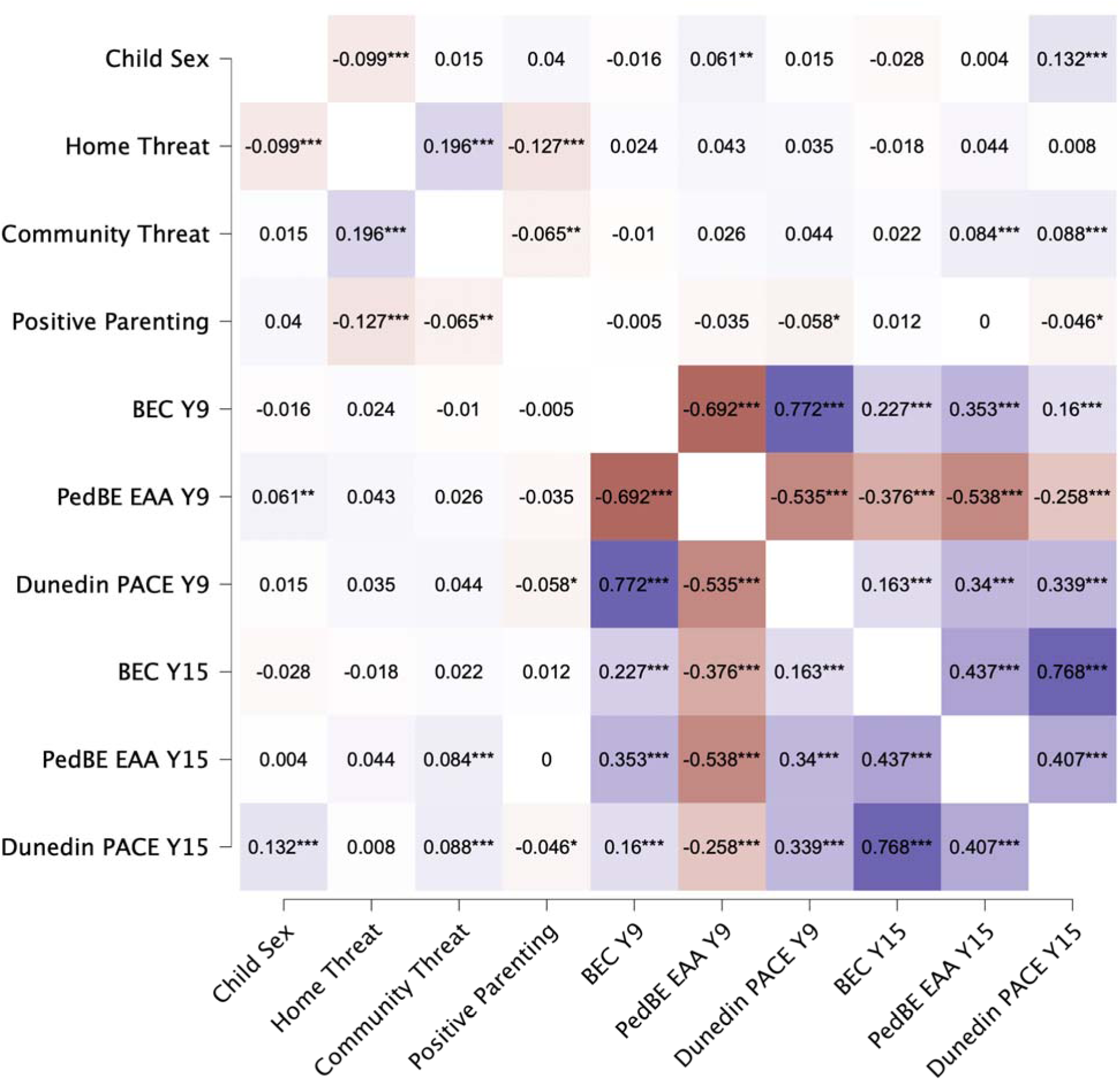
Bivariate correlations heatmap. Note. BEC = buccal epithelial cell proportion; EAA = epigenetic age acceleration; PACE = epigenetic pace of aging.

**Figure 3.**
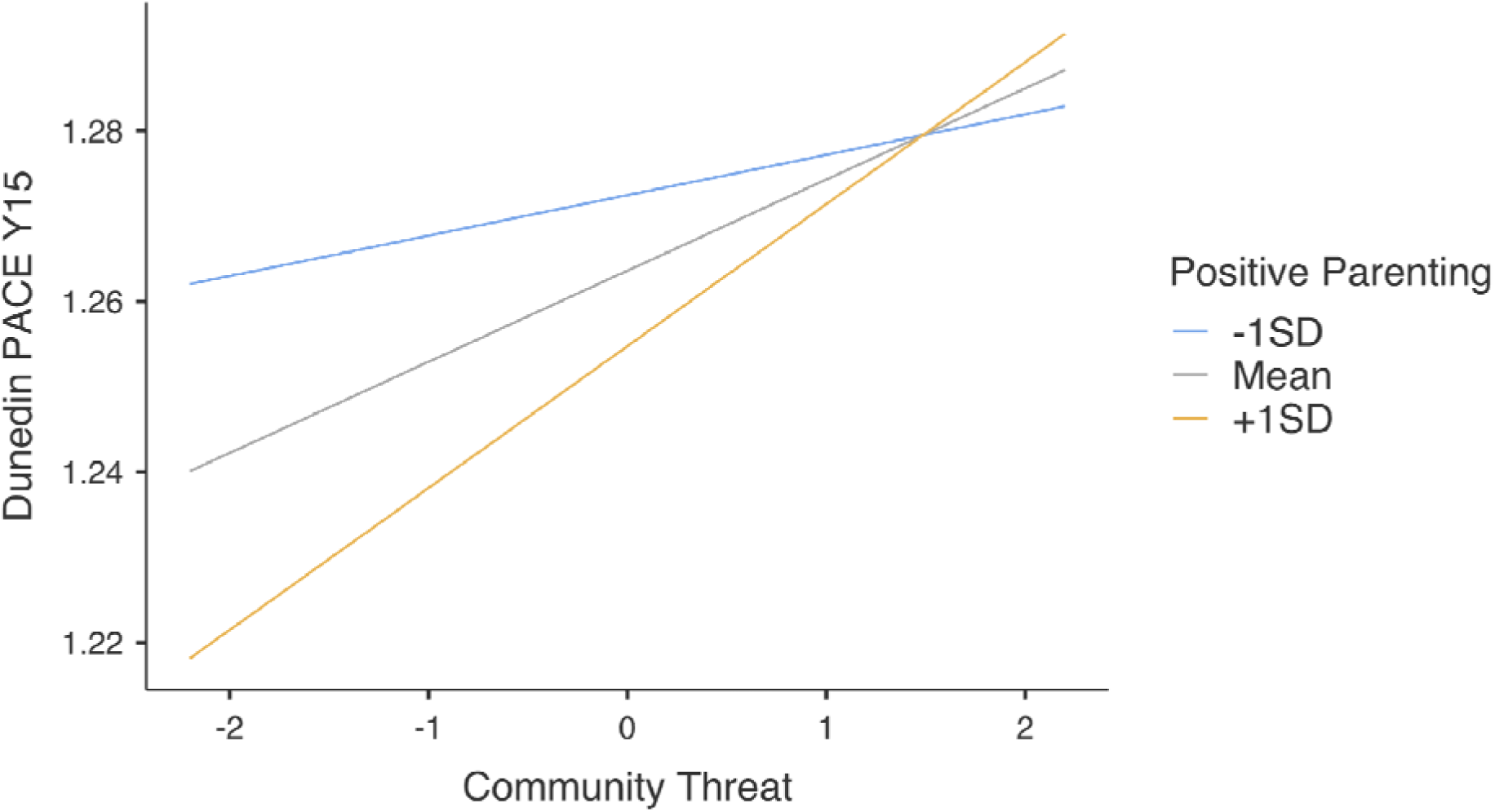
Moderation of community threat by positive parenting on epigenetic pace of aging. Note. Plotting points are estimated based on complete data (n = 1904) in Jamovi due to plotting limitations in Mplus.

## Discussion

Substantial research has established the long-term physical and mental health consequences of growing up in the face of adversity. However, far less is known about the biological mechanisms underlying these associations, particularly the modifiable protective factors that may buffer against the biological embedding of ELA. This longitudinal study addressed these gaps by examining whether positive parenting practices may buffer the impact of threat-based ELA within the home and the community on accelerated epigenetic aging in adolescence. Consistent with previous work (Hogan et al., 2024), greater childhood exposure to home– and community-based adversity was associated with accelerated epigenetic aging in adolescence. However, only community-based threat significantly predicted a faster pace of aging, as measured by DunedinPACE. Moreover, positive parenting buffered the effect of community-based threat on pace of aging, but this buffering was most evident under low to moderate levels of community-based threat. In contrast, positive parenting did not moderate the effects of home-based threat in the context of pace of aging, nor did it significantly interact with either form of threat to predict PedBE epigenetic age. These findings highlight the contextual complexity of how adversity and support systems interact to shape biological aging. Rather than acting as a universal buffer, positive parenting appears to have a conditional effect, reducing biological risk primarily when community-level stress is not overwhelming. This is consistent with prior findings suggesting that parenting may protect against mild to moderate stressors, but its effect is limited when adversity exceeds a certain threshold (Mendez et al., 2016). In this context, extreme community-based threat may overwhelm the protective capacity of even the most responsive caregivers, a phenomenon aligned with stress proliferation theory (Pearlin, 1999), which posits that high levels of stress can cascade across domains and undermine existing supports.

Situating these findings within Bronfenbrenner’s bioecological model (Bronfenbrenner & Ceci, 1994), our results underscore how mesosystem dynamics (e.g., interactions between levels) may either amplify or buffer longitudinal biological risk. Notably, the analyses demonstrate that adolescents who experienced both *low* community-based threat and *high* positive parenting early in life showed the *slowest* epigenetic aging. This finding suggests that risk often work in tandem rather than independently. For instance, these mesosystem interactions are nested within larger macrosystem forces—such as structural racism, socioeconomic status, access to resources and systemic inequality—which likely shape both parenting and community-level exposures (Stern et al., 2022). Multiple studies have found that White Americans show marginally slower biological aging than Black, Latinx, and Asian Americans which is a disparity theorized to reflect cumulative psychosocial and environmental stress (Brody et al., 2014; Farina et al., 2023; Raffington et al., 2023). These patterns reinforce the need for multilevel interventions that address not only parenting and community contexts but also the structural inequities embedded across broader socioecological systems which drive accelerated aging trajectories. Notably, in sensitivity analyses adjusting for family poverty ratio, the observed effects of threat-based adversity and its interaction with positive parenting on DunedinPACE remained significant, while no effects emerged for PedBE. These results suggest that the pathways linking threat exposure to biological aging are not fully explained by broader socioeconomic disadvantage but may reflect mechanisms specific to threat-related exposures.

From a Life History Theory (LHT) perspective, a faster pace of biological aging may represent an *adaptive calibration* to early environments characterized by threat and unpredictability (Belsky et al., 2012). In such contexts, accelerating development and prioritizing immediate survival may be biologically advantageous—even if these adaptations carry long-term health risks. This perspective highlights that biological systems are not passive victims of adversity, but rather active, responsive mechanisms influenced by environmental demands. However, the conditional effects of positive parenting observed in this study suggest that not all children exposed to adversity follow a uniformly accelerated trajectory. Instead, protective inputs like supportive caregiving may modulate or recalibrate overactive stress-response systems, shaping developmental timing in ways that reduce physiological wear and tear over time.

Our findings underscore the heterogeneity of outcomes following adversity and the importance of identifying when, in what contexts, and for whom protective factors are most effective. In particular, the present results suggest that positive parenting may buffer against the psychological consequences of ELA exposure even if it doesn’t undo the underlying biological adaptations. Future research should examine how multiple, interacting systems—from caregiving and schooling to neighborhoods and structural conditions—shape the biological embedding of stress. By investigating multiple risk and protective factors within specific environments, it could aid in facilitating the creation of better resources to help children and adolescents who have faced significant adversity. For example, DiMarzio et al. (2024) found that positive perceptions of school climate were associated with better psychosocial outcomes in adolescents who had experienced emotional maltreatment, highlighting the importance of ecological support beyond the home and the positive influence of a safe school environment. Together, this work suggests that resilience may be most robust when support is layered across ecological levels and contexts, even if individual-level adaptations persist.

Interestingly, our results revealed divergent patterns across two commonly used measures of epigenetic aging. While DunedinPACE was sensitive to both community-based threat and the moderating effect of positive parenting, PedBE accelerated age was predicted only by threat exposure, with no evidence of moderation by parenting. This divergence likely reflects both conceptual and biological differences between the two clocks. DunedinPACE estimates the pace of systemic aging across multiple physiological domains, whereas PedBE was developed to predict chronological age in children, based on DNA methylation patterns in buccal epithelial cells. Notably, the two measures do not share any CpG sites, despite both being biomarkers tied to age-related processes. These findings support the view that epigenetic clocks are not interchangeable but instead capture distinct pathways of biological development that may respond differentially to a multitude of environmental contexts.

Prior research aligns with this interpretation as multiple studies have identified unique differences between environmental interaction and biological aging. For example, Sullivan et al. (2023) found interactive effects between adversity and parenting on PedBE aging, while Merrill, Hogan, and colleagues (2024) identified main effects of positive parenting, regardless of adversity, on slowing down DunedinPACE. Together, these results suggest that epigenetic aging is multidimensional, and that developmental timing, tissue type, and the nature of adversity or support may each influence which biological pathways are most affected. By integrating multiple epigenetic clocks, each with different sensitivities, it could lead to a better understanding of how early experiences shape long-term health trajectories through underlying biological mechanisms. Given ongoing debates about how to define biological aging, what outcomes should be used to validate aging biomarkers (Herzog et al., 2024), and the limited focus on childhood and adolescence in this field (Raffington, 2024), future research must work toward a more unified framework to better identify biological factors that indicate future risk.

Finally, the absence of an interaction between positive parenting and home-based threat may be due to the unique dynamics of positive parenting. Research has shown that harsh and supportive parenting practices are not mutually exclusive and can exist simultaneously, which may create a complex and potentially contradictory environment for children (Parent et al., 2016). Such co-occurrence may dilute the protective effects of positive parenting, as the stress induced by threat-based parenting practices (e.g., corporal punishment, hostility) could undermine the benefits of warmth and responsiveness. This aligns with broader theories suggesting that the protective capacity of parenting practices can be context-dependent and may vary based on the intensity, duration, and type of adversity experienced (Mclaughlin et al., 2014). Future research is needed to examine how these parenting practices interact dynamically over time and whether certain combinations of supportive and harsh parenting can have differential effects on biological aging outcomes.

## Strengths and Limitations

This study has several limitations that should be considered when interpreting findings. First, the initial sample was disproportionately skewed to non-marital births in economically disadvantaged families, and predominantly from historically minoritized racial or ethnic backgrounds living in urban areas. Thus, results may not represent the larger United States population or other counties, rural neighborhoods, and a wide array of racial/ethnic groups or genetic ancestry backgrounds. However, social epigenetic research has been predominantly conducted with White European ancestry samples, and increased research with populations who experience health disparities is needed to better understand and address the drivers of health disparities and inform the development of effective intervention and prevention programs among various underserved populations (Gillman et al., 2024).

Further, community-based threat was measured using objective crime rate data from the neighborhoods in which children and their parents resided during the first nine years of life, rather than through direct, self-reported experiences of community violence. While the use of objective crime data may provide important contextual information, it may not accurately capture the participants’ *subjective* or *perceived* experiences of threat. This is an important distinction given that perceived threat is often more predictive of psychological and biological outcomes than potential exposure alone. Nonetheless, the use of objective indicators still offers valuable insight into the ambient threat level of a child’s environment and suggests that even potential exposure to violence may shape developmental trajectories through both direct and indirect pathways (Colich et al., 2020; McLaughlin et al., 2014).

While we controlled baseline epigenetic aging at age 9, we were limited by the absence of epigenetic data prior to that point and did not include concurrent measures of adversity or parenting during adolescence. As a result, our models cannot fully capture the dynamic, reciprocal nature of these processes across development. Future research that incorporates repeated assessments of adversity, parenting, and epigenetic outcomes across multiple developmental stages—or employs experimental designs—will be better positioned to establish temporal ordering, detect developmental changes, and draw stronger causal inferences. Additionally, because DNA methylation in this study was derived from saliva, and most epigenetic clocks were originally developed using other tissue types (e.g., blood, buccal cells), future work should assess the robustness of these associations across multiple biological samples to enhance generalizability.

Despite its limitations, the study demonstrates several key strengths in its design and statistical implementation that also should be considered. First, the longitudinal design and multi-method assessment (i.e., parent self-reports, geocoded neighborhood data, observed parenting, and biomarkers) spanning multiple waves from childhood to adolescence strengthens confidence in findings. Further, we explored multiple domains of threat-based ELA, and findings differed based on the environmental context in which they were experienced. This may aid in the development of new intervention strategies that are tailored to specific socio-ecological contexts with the aim of minimizing the biological embedding of adversity across subsequent generations. Lastly, this study explored the role of positive parenting in buffering the impact of adversity, which contributes to the growing, but still limited, literature studying mechanisms of health longevity and resilience across child and adolescent development (Merrill, Konwar, et al., 2024).

## Conclusion

In summary, our findings contribute to the growing understanding of how early life environmental contexts shape biological aging across the lifespan. Specifically, they underscore the need for continued research to clarify the multidimensional pathways linking ELA to accelerated epigenetic aging and to identify protective factors—such as parenting practices—that may buffer these effects. Our results suggest that either high levels of community-based threat or low levels of positive parenting are sufficient to accelerate biological aging. Conversely, the co-occurrence of high positive parenting and low community-based threat is associated with the slowest pace of epigenetic aging. These results highlight that biological resilience is shaped through the interplay between multiple contextual systems and the individual—not by individual traits alone. Importantly, our findings were robust to adjustment for family poverty, indicating that threat-related exposures contribute uniquely to biological aging beyond the effects of socioeconomic disadvantage. Moving forward, research should explore how risk and protective factors operate across developmental stages and ecological contexts, and how multi-level interventions can most effectively disrupt the biological embedding of early life adversity. Such work is essential to inform public health initiatives that aim to reduce disparities in aging and health across the lifespan.

## Data Availability

The current study uses publicly available data from the Future of Families and Child Wellbeing Study (https://ffcws.princeton.edu/).

## Acknowledgments

Research reported in this publication was supported by the Eunice Kennedy Shriver National Institute of Child Health and Human Development (NICHD) of the National Institutes of Health under award numbers R01HD036916, R01HD039135, and R01HD040421, as well as a consortium of private foundations. AH was supported by NICHD T32HD101392 (Stroud & Tyrka, MPIs). KD was supported by NICHD F31HD106768. SD was supported by the National Science Foundation Graduate Research Fellowship under Grant No. 2146759. DMR is supported by NHLBI 1K01HL169495 and NIGMS P20GM139767 (Stroud, Laura). JP was partially supported by the Rhode Island Institutional Development Award (IDeA) Network of Biomedical Research Excellence from the National Institute of General Medical Sciences of the National Institutes of Health under grant number P20GM103430. NICHD L40HD103048-03 and NIMHD R01MD015401 also supported this work. The content is solely the responsibility of the authors and does not necessarily represent the official views of the National Institutes of Health.

## Open Science Declaration

The data used in this study (Future of Families and Child Wellbeing Study) are publicly available at https://ffcws.princeton.edu/documentation. The analyses were not preregistered. The analytic code used in this study is available from the corresponding author upon reasonable request. No new materials were created for this study.

