## Supplemental Appendix for "Community Threat, Positive Parenting, and Accelerated Epigenetic Aging: Longitudinal Links from Childhood to Adolescence"

Table S1 – Descriptives for raw indicator variables used in factor models……………………….. 2

Table S2 – CFA model results for the threat variables………………………………………………. 4

Table S3 – CFA model results for parenting………………………………………………………….. 6

Figure S1 – CFA results for the threat model………………………………………………………… 7

Figure S2 – CFA results for the parenting model……………………………………………………. 8

**Table S1** – Descriptives for raw indicator variables used in factor models

| **Construct** | **Year** | **Variable Name** | **Question** | **Respondent** | **Coding** | **M / %** | **SD** |
| --- | --- | --- | --- | --- | --- | --- | --- |
| Parenting | 3 | o3t7 | Parent spontaneously praises child at least twice | Interviewer | 0= Did not occur (age 9 reverse coded)  1= Did occur (age 9 reverse coded) | 82.90% |  |
|  |  | o3t8 | Parent’s voice conveys positive feelings toward child |  |  | 93.80% |  |
|  |  | o3t9 | Parent caressed or kissed child at least once |  |  | 78.60% |  |
|  | 5 | o4t5 | Parent spontaneously praised child's behavior or qualities twice |  |  | 68.70% |  |
|  |  | o4t6 | Parent used some term of endearment twice or more during visit |  |  | 61.30% |  |
|  |  | o4t7 | Parent's voice conveyed positive feelings when speaking of or to child |  |  | 93.10% |  |
|  |  | o4t8 | Parent caressed or kissed child at least once |  |  | 58.60% |  |
|  | 9 | o5e3 | Parent/PCG encouraged child to contribute to conversation |  |  | 69.80% |  |
|  |  | o5e4 | Parent/PCG helped child demonstrate achievement or skill during visit |  |  | 59.20% |  |
|  |  | o5e5 | Parent/PCG used term of endearment twice during visit |  |  | 48.50% |  |
|  |  | o5e6 | Parent/PCG's voice conveyed positive feelings |  |  | 92.20% |  |
|  |  | o5e7 | Parent/PCG caressed, kissed, or cuddled child once during visit |  |  | 31.00% |  |
| Home Threat | 3 | p3j4 | Past year, times PCG hit child on the bottom with belt or hard object | PCG | 0 = Never  1 = Once  2 = Twice  3 = 3-5 times  4 = 6-10 times  5 = 11-20 times  6 = More than 20 times  7 = Yes, but not in the past year (recoded as 0) | 0.8 | 1.636 |
|  |  | p3j6 | Past year, times PCG shouted, yelled, or screamed at child |  |  | 3.46 | 2.145 |
|  |  | p3j7 | Past year, times PCG spanked child on bottom with bare hand |  |  | 2.72 | 2.122 |
|  |  | p3j8 | Past year, times PCG swore or cursed at child |  |  | 0.56 | 1.359 |
|  |  | p3j10 | Past year, times PCG threatened to spank/hit child but did not |  |  | 3.63 | 2.362 |
|  |  | p3j11 | Past year, times PCG slapped child on the hand, arm, or leg |  |  | 2.27 | 2.178 |
|  | 5 | p4g4 | Past year, times PCG hit child on the bottom with belt or hard object | PCG | 0 = Never  1 = Once  2 = Twice  3 = 3-5 times  4 = 6-10 times  5 = 11-20 times  6 = More than 20 times  7 = Yes, but not in the past year (recoded as 0) | 0.98 | 1.655 |
|  |  | p4g6 | Past year, times PCG shouted, yelled, or screamed at child |  |  | 3.75 | 2.02 |
|  |  | p4g7 | Past year, times PCG spanked child on bottom with bare hand |  |  | 2.3 | 2.04 |
|  |  | p4g8 | Past year, times PCG swore or cursed at child |  |  | 0.77 | 1.529 |
|  |  | p4g10 | Past year, times PCG threatened to spank/hit child but did not |  |  | 3.63 | 2.318 |
|  |  | p4g11 | Past year, times PCG slapped child on the hand, arm, or leg |  |  | 1.79 | 2.001 |
|  | 9 | p5q1d | Parent has hit child on bottom with something like brush or other hard object | PCG | 1 = Once  2 = Twice  3 = 3-5 times  4 = 6-10 times  5 = 11-20 times  6 = More than 20 times  7 = Yes, but not in the past year (recoded as 0)  8 = Never (recoded as 0) | 0.85 | 1.381 |
|  |  | p5q1f | Parent has shouted, yelled or screamed at child |  |  | 3.1 | 1.907 |
|  |  | p5q1g | Parent has spanked child on bottom with bare hand |  |  | 1.22 | 1.559 |
|  |  | p5q1h | Parent has swore or cursed at child |  |  | 0.87 | 1.449 |
|  |  | p5q1j | Parent has threatened to spank or hit child but did not actually do it |  |  | 2.54 | 2.199 |
|  |  | p5q1k | Parent has slapped child on hand, arm or leg |  |  | 1.09 | 1.541 |
| Community Threat | 3 | rg3ucr_mallrt | Mother county total crime rate (per 100k people), Wave 3 interview | X | X | 5339.32 | 1880.57 |
|  | 5 | rg4ucr_mallrt | Mother county total crime rate (per 100k people), Wave 4 interview |  |  | 5128.75 | 1829.26 |
|  | 9 | rg5ucr_mallrt | Mother county total crime rate (per 100k people), Wave 5 interview |  |  | 4556.56 | 1657.06 |

**Table S2** – CFA model results for the threat variables

| **Section** | **Variable** | **Estimate** | **S.E.** | **Est./S.E.** | **P-Value** |
| --- | --- | --- | --- | --- | --- |
| Y3_HTHR BY | P3J6 | 1.000 | 0.000 | 999.000 | 999.000 |
| Y3_HTHR BY | P3J10 | 1.139 | 0.027 | 41.675 | 0.000 |
| Y3_HTHR BY | P3J8 | 0.891 | 0.033 | 26.621 | 0.000 |
| Y3_HTHR BY | P3J7 | 1.181 | 0.026 | 45.541 | 0.000 |
| Y3_HTHR BY | P3J4 | 0.886 | 0.032 | 27.543 | 0.000 |
| Y3_HTHR BY | P3J11 | 1.159 | 0.026 | 45.247 | 0.000 |
| Y5_HTHR BY | P4G6 | 1.000 | 0.000 | 999.000 | 999.000 |
| Y5_HTHR BY | P4G10 | 1.161 | 0.034 | 34.363 | 0.000 |
| Y5_HTHR BY | P4G8 | 0.949 | 0.035 | 27.445 | 0.000 |
| Y5_HTHR BY | P4G7 | 1.111 | 0.032 | 34.673 | 0.000 |
| Y5_HTHR BY | P4G4 | 0.953 | 0.034 | 27.956 | 0.000 |
| Y5_HTHR BY | P4G11 | 1.124 | 0.032 | 35.294 | 0.000 |
| Y9_HTHR BY | P5Q1F | 1.000 | 0.000 | 999.000 | 999.000 |
| Y9_HTHR BY | P5Q1J | 1.186 | 0.027 | 44.610 | 0.000 |
| Y9_HTHR BY | P5Q1H | 0.963 | 0.027 | 35.161 | 0.000 |
| Y9_HTHR BY | P5Q1G | 0.944 | 0.026 | 35.757 | 0.000 |
| Y9_HTHR BY | P5Q1D | 1.068 | 0.027 | 39.493 | 0.000 |
| Y9_HTHR BY | P5Q1K | 1.049 | 0.026 | 40.018 | 0.000 |
| Y359GEO BY | ZRG3_ALL | 1.000 | 0.000 | 999.000 | 999.000 |
| Y359GEO BY | ZRG4_ALL | 1.037 | 0.032 | 32.803 | 0.000 |
| Y359GEO BY | ZRG5_ALL | 0.865 | 0.039 | 22.303 | 0.000 |
| Y349HTR BY | Y3_HTHR | 1.000 | 0.000 | 999.000 | 999.000 |
| Y349HTR BY | Y5_HTHR | 1.125 | 0.052 | 21.663 | 0.000 |
| Y349HTR BY | Y9_HTHR | 0.939 | 0.040 | 23.701 | 0.000 |
| Y3_HTHR WITH | Y5_HTHR | 0.000 | 0.000 | 999.000 | 999.000 |
| Y3_HTHR WITH | Y9_HTHR | 0.000 | 0.000 | 999.000 | 999.000 |
| Y5_HTHR WITH | Y9_HTHR | 0.000 | 0.000 | 999.000 | 999.000 |
| Y359GEO WITH | Y349HTR | 0.086 | 0.010 | 8.743 | 0.000 |
| P3J6 WITH | P4G6 | 0.179 | 0.015 | 12.312 | 0.000 |
| P3J6 WITH | P5Q1F | 0.154 | 0.015 | 10.237 | 0.000 |
| P4G6 WITH | P5Q1F | 0.238 | 0.015 | 16.254 | 0.000 |
| P3J10 WITH | P4G10 | 0.136 | 0.015 | 9.252 | 0.000 |
| P3J10 WITH | P5Q1J | 0.114 | 0.015 | 7.812 | 0.000 |
| P4G10 WITH | P5Q1J | 0.128 | 0.015 | 8.676 | 0.000 |
| P3J8 WITH | P4G8 | 0.306 | 0.023 | 13.563 | 0.000 |
| P3J8 WITH | P5Q1H | 0.247 | 0.023 | 10.860 | 0.000 |
| P4G8 WITH | P5Q1H | 0.272 | 0.020 | 13.310 | 0.000 |
| P3J7 WITH | P4G7 | 0.138 | 0.013 | 10.383 | 0.000 |
| P3J7 WITH | P5Q1G | 0.079 | 0.016 | 5.022 | 0.000 |
| P4G7 WITH | P5Q1G | 0.124 | 0.017 | 7.319 | 0.000 |
| P5Q1G WITH | P5Q1K | 0.159 | 0.015 | 10.793 | 0.000 |
| P3J4 WITH | P4G4 | 0.300 | 0.021 | 14.334 | 0.000 |
| P3J4 WITH | P5Q1D | 0.198 | 0.022 | 9.049 | 0.000 |
| P4G4 WITH | P5Q1D | 0.273 | 0.018 | 14.814 | 0.000 |
| P3J11 WITH | P4G11 | 0.084 | 0.015 | 5.631 | 0.000 |
| P3J11 WITH | P5Q1K | 0.085 | 0.017 | 4.913 | 0.000 |
| P4G11 WITH | P5Q1K | 0.108 | 0.017 | 6.458 | 0.000 |
| Intercepts | ZRG3_ALL | 0.000 | 0.016 | 0.000 | 1.000 |
| Intercepts | ZRG4_ALL | 0.000 | 0.016 | 0.000 | 1.000 |
| Intercepts | ZRG5_ALL | 0.000 | 0.017 | 0.000 | 1.000 |
| Variances | Y349HTR | 0.261 | 0.017 | 15.762 | 0.000 |
| Variances | Y359GEO | 0.773 | 0.037 | 21.083 | 0.000 |
| Residual Variances | ZRG3_ALL | 0.227 | 0.026 | 8.841 | 0.000 |
| Residual Variances | ZRG4_ALL | 0.168 | 0.023 | 7.395 | 0.000 |
| Residual Variances | ZRG5_ALL | 0.421 | 0.024 | 17.918 | 0.000 |
| Residual Variances | Y3_HTHR | 0.201 | 0.013 | 14.971 | 0.000 |
| Residual Variances | Y5_HTHR | 0.106 | 0.014 | 7.519 | 0.000 |
| Residual Variances | Y9_HTHR | 0.263 | 0.014 | 18.244 | 0.000 |

**Table S3** – CFA model results for parenting

| **Factor** | **Variable** | **Estimate** | **S.E.** | **Est./S.E.** | **P-Value** |
| --- | --- | --- | --- | --- | --- |
| Y3POSPAR BY | O3T7 | 1.000 | 0.000 | 999.000 | 999.000 |
| Y3POSPAR BY | O3T8 | 0.931 | 0.051 | 18.281 | 0.000 |
| Y3POSPAR BY | O3T9 | 0.817 | 0.046 | 17.750 | 0.000 |
| Y5POSPAR BY | O4T5 | 1.000 | 0.000 | 999.000 | 999.000 |
| Y5POSPAR BY | O4T6 | 0.941 | 0.035 | 27.024 | 0.000 |
| Y5POSPAR BY | O4T7 | 1.000 | 0.043 | 23.281 | 0.000 |
| Y5POSPAR BY | O4T8 | 0.868 | 0.034 | 25.899 | 0.000 |
| Y9POSPAR BY | O5E3 | 1.000 | 0.000 | 999.000 | 999.000 |
| Y9POSPAR BY | O5E4 | 1.012 | 0.035 | 28.961 | 0.000 |
| Y9POSPAR BY | O5E5 | 0.979 | 0.033 | 29.768 | 0.000 |
| Y9POSPAR BY | O5E6 | 0.994 | 0.040 | 24.776 | 0.000 |
| Y9POSPAR BY | O5E7 | 0.879 | 0.034 | 26.185 | 0.000 |
| Y359POS BY | Y3POSPAR | 1.000 | 0.000 | 999.000 | 999.000 |
| Y359POS BY | Y5POSPAR | 1.323 | 0.316 | 4.190 | 0.000 |
| Y359POS BY | Y9POSPAR | 0.625 | 0.110 | 5.689 | 0.000 |
| Y3POSPAR WITH | Y5POSPAR | 0.000 | 0.000 | 999.000 | 999.000 |
| Y3POSPAR WITH | Y9POSPAR | 0.000 | 0.000 | 999.000 | 999.000 |
| Y5POSPAR WITH | Y9POSPAR | 0.000 | 0.000 | 999.000 | 999.000 |
| Y5POSPAR WITH | Y9POSPAR | 0.000 | 0.000 | 999.000 | 999.000 |
| Variances | Y359POS | 0.213 | 0.059 | 3.619 | 0.000 |
| Residual Variances | Y3POSPAR | 0.671 | 0.074 | 9.097 | 0.000 |
| Residual Variances | Y5POSPAR | 0.382 | 0.099 | 3.877 | 0.000 |
| Residual Variances | Y9POSPAR | 0.519 | 0.033 | 15.641 | 0.000 |

**Figure S1** – CFA results for the threat model

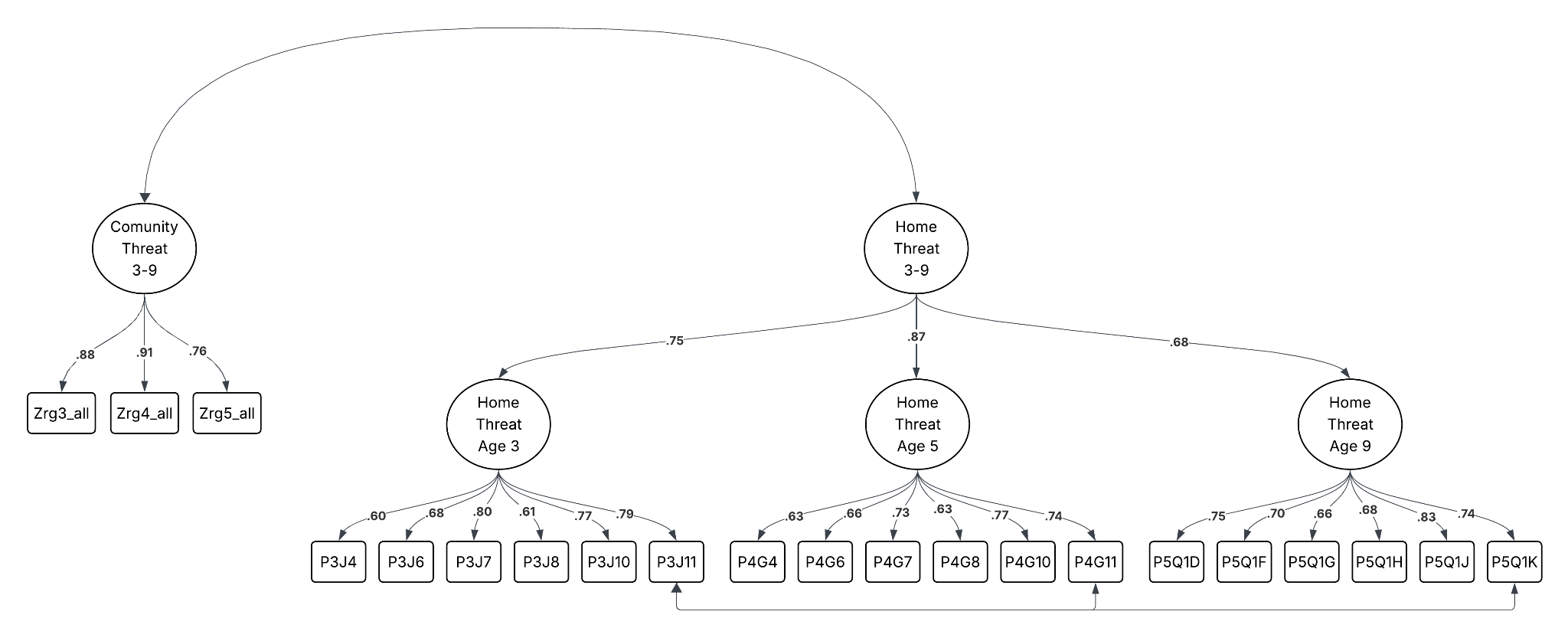

Note. One example set of indicator residual covariances is depicted for demonstration. See Table S1 for complete results

**Figure S2** – CFA results for the parenting model

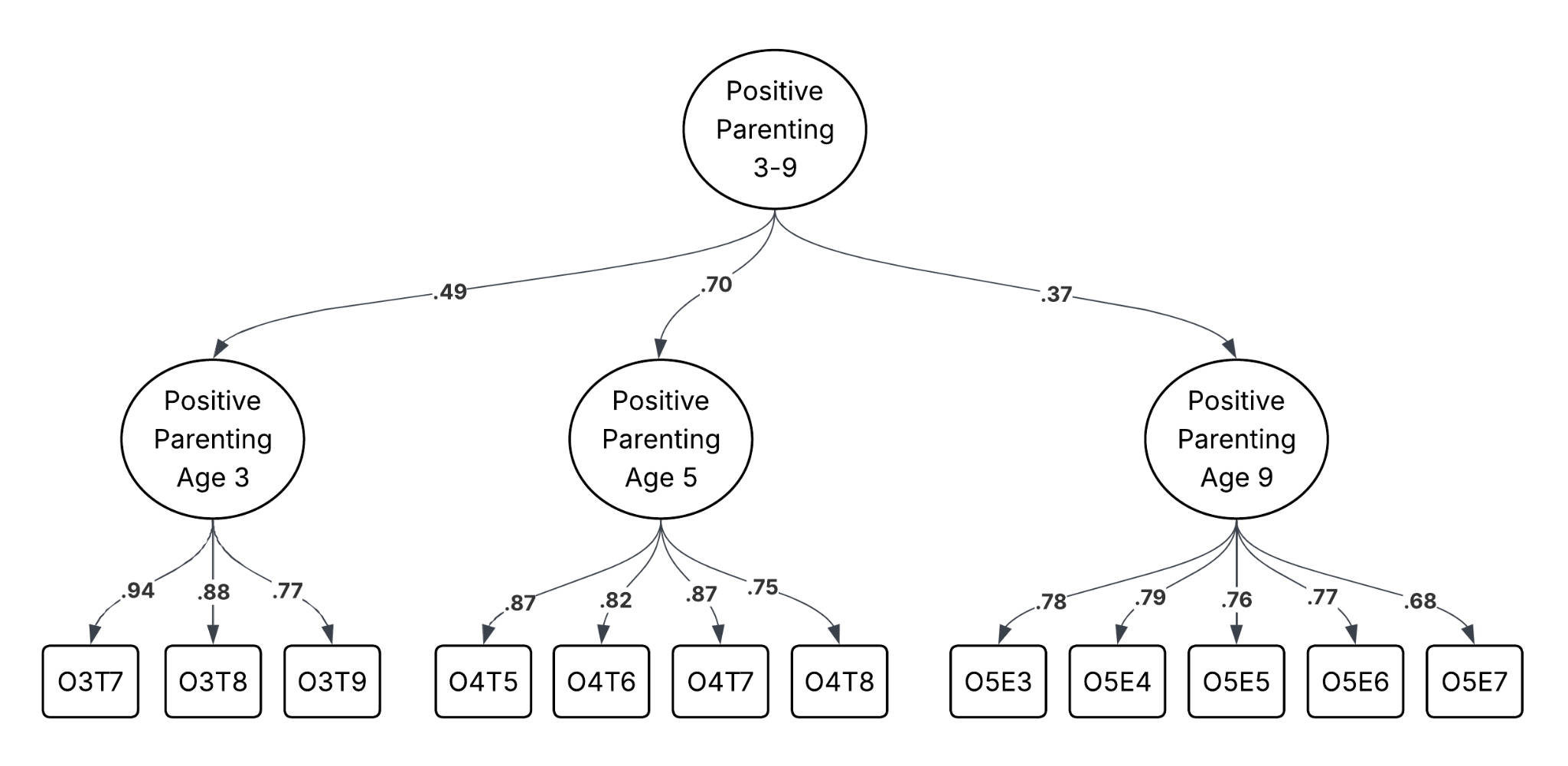
